# The concurrent development of psychological distress, heavy episodic drinking, and daily smoking from adolescence to midlife in two cohorts

**DOI:** 10.1101/2024.03.06.24303863

**Authors:** Noora Berg, Maarit Piirtola, Mauri Marttunen, Antti Latvala, Olli Kiviruusu

**Affiliations:** Department of Public Health and Welfare, Finnish Institute for Health and Welfare, P.O. Box 30, 00271, Helsinki, Finland; Institute for Molecular Medicine Finland, HiLIFE, University of Helsinki, P.O. Box 20, 00014, Helsinki, Finland; UKK Institute for Health Promotion Research, Kaupinpuistonkatu 1, 33500Tampere, Finland; Adolescent Psychiatry, Helsinki University and Helsinki University Hospital, P.O. Box 660, 00029 HUS, Helsinki, Finland; Institute of Criminology and Legal Policy, University of Helsinki, P.O. Box 16, 0014, Helsinki, Finland

**Keywords:** alcohol use, heavy episodic drinking, life course, mental health, psychological distress, smoking, tobacco use

## Abstract

**Background:** Heavy substance use, such as alcohol and tobacco use, is more prevalent among people with poor mental health. Despite the well-established correlation between substance use and mental health, the development of this association over time is unclear. The aim of this study was to examine the development and co-occurrence of psychological distress (mainly depressive and anxiety symptoms), heavy episodic drinking (HED) and daily smoking from adolescence to adulthood.

**Methods:** Two prospective longitudinal studies, the Stress, Development and Mental Health Study (TAM, N=2194) and the FinnTwin16 Study (N=5563), have followed participants from adolescence to adulthood (TAM ages 16-52, FinnTwin16 ages 16-35) using survey questionnaires. Latent class analysis was used to obtain joint trajectories of distress, HED and daily smoking.

**Results:** This study identified several different patterns of how HED, daily smoking and psychological distress cluster across time from adolescence to adulthood. In both examined cohorts, distinct groups of i) low levels of all three examined health concerns, ii) high levels of all three concerns, and iii) high distress and low-to-moderate substance use were found. In the older TAM cohort with longer follow-up time, a group indicating iv) increasing HED and a group indicating v) all on average level, with a peak in smoking, were also found.

**Conclusions:** Found variations set requirements for substance use and mental health services to target interventions for different groups to address all three major public health problems.

## Background

Poor mental health and high consumption of alcohol and tobacco are significant public health concerns (1,2). Heavy alcohol use and smoking are more prevalent among people with poor mental health (3,4). Although the mechanisms underlying this association, including the direction of possible causal effects between mental health and substance use, are not fully understood, both substance use and poor mental health have been identified as risk factors for each other (3,5–9). High use of alcohol and tobacco is also a contributing factor to lower life expectancy among people with mental disorders (10). To address this cumulative burden of poor mental health and substance use, we need new perspectives on this well-acknowledged association between mental health and substance use. Life course theory (LCT) is an interdisciplinary framework that can combine individual, contextual and temporal aspects to better understand the association between poor mental health and heavy substance use (11).

### Age curves of alcohol use, smoking and psychological distress

There is a growing number of studies examining stability and changes in psychological distress/symptoms/disorders and substance use over time. At the population level, the normative age curve for substance use typically increases in adolescence and young adulthood and then decreases (alcohol) or stabilizes (tobacco) thereafter (12,13). The reasons for use vary by age: experiments are common in adolescence and young adulthood whereas the use of substances to regulate stress and emotions increases the risk of e.g. adult alcohol use disorder (AUD) (14). Regarding the association of increasing age with mental health, there is more variation in the literature depending on whether under investigation has been e.g., symptoms vs. diagnosis, prevalence vs. incidence, depressive symptoms vs. more general psychological distress, cross-sectional vs. birth cohort vs. cohort sequential study design. Nevertheless, the incidence of depression has typically been found to increase from childhood to adolescence and to be at highest in young adulthood (15). Correspondingly, studies have quite consistently shown that the prevalence of depression increases from childhood to young adulthood, and in Western countries decreases after young adulthood, but there are differing views on whether the prevalence decreases or stabilizes/increases in other regions (16,17). Regarding anxiety, the evidence is less clear; in general, studies have shown an increase in the prevalence of anxiety disorders from adolescence to adulthood and a decrease until old age, but there is variation in the type of symptoms/disorders (increase in e.g. panic disorder and a decrease in social phobia from adolescence to early adulthood)(7,18–21). Genetic effects have been found to explain some but not all of the variation in the age curve (22). However, both psychological distress/depression and anxiety symptoms/disorders are common worldwide and cause burdens on health and wellbeing (23–25).

### Heterogeneity in the age curves (trajectory studies)

An increasing number of studies have examined substance use and mental health trajectories and highlighted the heterogeneity in the age curve (26–28). However, only a few studies have examined the joint trajectories of all three aspects: mental health and alcohol and tobacco use. Simultaneous modelling the developmental course of several distinct but related outcomes provides a rich yet easily comprehended statistical summary of the developmental linkages between the outcomes of interest (29). Examining heterogeneity in the joint trajectories of mental health and substance use during the life course is important for identifying potential subgroups of people with differing, age-specific needs for prevention and treatment. Lee et al. (30) examined joint trajectories of alcohol use, tobacco use and depressive symptoms from adolescence to adulthood (aged 14-29) among African Americans and Puerto Ricans born in the mid-1970s in the Harlem Longitudinal Development Study. They found 5 trajectory groups: 1) moderate alcohol use, high tobacco use, and high depressive symptoms (12%); 2) moderate alcohol use, high tobacco use, and low depressive symptoms (26%); 3) moderate alcohol use, low tobacco use, and low depressive symptoms (18%); 4) low alcohol use, no tobacco use, and high depressive symptoms (11%); and 5) low alcohol use, no tobacco use, and low depressive symptoms (33%).

Although triple trajectory studies are rare, there is some evidence of intertwined joint pathways from studies that have modelled two out of the three phenomena longitudinally. Regarding studies on alcohol use and mental health, an Australian age-period-cohort study concluded that heavy episodic drinking (HED) (≥4 drinks in a row) and psychological distress remained positively related across the lifespan (31). A Finnish study examining whether the development of psychological symptoms differed between persons with different developmental paths of HED from adolescence to midlife revealed that the greater the HED trajectory indicated frequent HED, the higher was the level of symptoms throughout the follow-up (32).

A 30-year follow-up study examining smoking and depressive symptoms from young adulthood to middle age revealed, that the association seems more robust among men than among women and that especially men who smoke persistently from young adulthood to middle age have an increased risk of high depressive symptoms trajectory (33). Another longitudinal study examined tobacco use and mental health with a 10-year follow-up and found that lifetime stress and anxiety disorders were more strongly associated with smoking trajectories (compared to nonsmoking trajectory) (34).

The association between alcohol and tobacco use is positive and they are intertwined at physiological, psychological and social levels (35,36). Many studies have examined joint or parallel trajectories of alcohol and tobacco use and found them to be positively correlated through the life course, although in most studies follow-up occurs or ends in young adulthood, at the peak of substance use (37–40). There are also studies modelling three components simultaneously, but instead of mental health the third component has often been marijuana use (41).

### Sociodemographic characteristics

It is important to understand the individual characteristics (e.g. sex) and early background (e.g. socioeconomic position) of the developmental paths of mental health and substance use to better target interventions preventing comorbidity and continuity of these issues.

### Sex

It is well known that men generally use alcohol and tobacco in larger amounts and more often than women, while women usually report more psychological distress (42,43). It has been suggested that this sex discrepancy in substance use and distress symptoms reflects gendered responses to negative affect (44). Internalizing symptoms reflect problems in emotion regulation and are more typical in women and externalizing symptoms reflect problems in behavior control and have been seen as more typical in men (43,45). Whether longitudinal pathways, i.e., the longitudinal linkages between substance use and mental health, also differ by sex is currently unclear.

### Socioeconomic background

According to a study by Lee et al. (30) with approximately 20 years of follow-up, the trajectory group with low alcohol use, no tobacco use and low depressive symptoms had the highest educational level, and the group with moderate alcohol use, high tobacco use and high depressive symptoms had the lowest. Overall, research on socioeconomic differences in substance use and mental health has concluded that i) people with higher SEP may consume similar or greater amounts of alcohol than people with lower SEP, although the latter group is more affected by negative alcohol-related consequences (46); ii) tobacco use is more common among lower SEP groups (47); and iii) socioeconomically disadvantaged people are more likely to develop mental health problems, although there is variation in the types of mental health problems (48).

### Summary

Few studies have examined the longitudinal associations between alcohol and tobacco use and mental health simultaneously, or how they fluctuate from adolescence to midlife. The co-occurrence of both mental health and substance use problems is associated with a more severe course of illness and present challenges for prevention and treatment strategies (49).

Most previous studies have focused on adolescence and young adulthood, a time period of accelerated substance use and psychological distress and have not considered the time period of the stabilizing or decreasing substance use later in life. There is a lack of studies that have longitudinally modelled both mental health and substance use using a life course perspective that spans several different life phases. Our study aims to address these gaps in the literature by considering both the developmental course and co-occurrence of these phenomena. By examining the interrelated longitudinal linkage between three different outcomes i.e. HED, smoking status and mental health, it is possible to reach a more comprehensive multidimensional view (50). This developmental approach provides information that helps to design interventions for the primary prevention of secondary health problems and to better tailor treatment services based on individual needs.

## Methods

### Aim

By integrating individual, contextual and temporal aspects and using the life course perspective as the framework this study aims to examine the development and co-occurrence of psychological distress, smoking and heavy episodic drinking from adolescence to midlife.

The specific research questions were as follows:

1. What kind of trajectory groups of concurrent heavy episodic drinking, smoking, and psychological distress from adolescence to midlife can be identified?
2. How are sociodemographic factors (sex, adolescent family structure and parental socioeconomic background) associated with trajectory group membership?

Furthermore, the present study investigated whether the combined trajectory classes of psychological distress and substance use were found in two different datasets with different age cohorts.

Based on previous research, we expected to find a rather large group with low levels of distress and both HED and smoking and a group with high levels of distress and both substances. Regarding other possible groups, we did not set specific hypothesis. We expected women to be assigned more to the distress-oriented trajectory groups and men and people in the low SEP group to be assigned more to the substance use oriented trajectory groups.

### Study population

This is a prospective multicohort study based on two longitudinal datasets from Finland that have followed participants from adolescence to adulthood. The studies are the ‘Stress, development and mental health’ (TAM) (N=2194) (51) and the ‘FinnTwin16’ (N=5563) (52).

The TAM consists of pupils (N=2194) who attended the last year of compulsory school in 1983 in Tampere, a city in southern Finland. The participants completed questionnaires at 16, 22, 32, 42, and 52 years of age (in 1983, 1989, 1999, 2009, and 2019). FinnTwin16 is a population-based study of five consecutive birth cohorts (1975–1979) of Finnish twins (N= 5563). Participants completed questionnaires at ages 16, 25, and 35 (in 1991-1995, 2000-2002, and 2010-2012).

In general, the participation rates were high. In the TAM, 53.0% of the original 16-year-old participants participated at age 52. In FinnTwin16, of those who were contacted, 72.0% participated at age 35.

### Measures

#### Psychological distress

Psychological distress included mainly depressive and anxiety symptoms. In the TAM the continuous measure was constructed from a 17-item psychosomatic symptoms checklist using seven items (on a scale from 0 to 3) indicative of depression and anxiety (lack of energy, sleeping difficulties, nightmares, fatigue, irritability, loss of appetite, and nervousness/anxiety). The measure has been used earlier and described in more detail in Pelkonen et al. (53). In FinnTwin16, a score on a symptom list of 4 items (tension or nervousness, fatigue, irritability, sleeping disorders) was used at age 16, and the General Health Questionnaire (GHQ-12) (54) was used at ages 25 and 35.

In the analyses, all symptom measures were standardized.

#### Heavy episodic drinking

Heavy episodic drinking (HED) was classified as i) having ≥5/6 drinks on one occasion or being drunk monthly or more often and ii) drinking the amount/being drunk more seldom or not at all. In the TAM cohort, HED was measured as the frequency of intoxication (16 and 22 years) and having six or more drinks in a row (32, 42 and 52 years). At age 16, the HED category included those who reported being drunk at least four times during the school term (on average once a month). At age 22, the HED category included those who reported heavy drunkenness at least once a month. At ages 32, 42, and 52, HED was based on the third question in the Alcohol Use Disorders Identification Test (AUDIT) (55); those who reported having six or more drinks in a session at least once a month were classified as heavy drinkers.

In the FinnTwin16 study, HED was measured as the frequency of intoxication (16 years) and having five or more drinks in a row (25 and 35 years). At age 16, the HED category included those who reported heavy drunkenness at least once a month. At ages 25 and 35, the HED category included those who reported having five or more bottles of beer, more than a bottle of wine or more than half a bottle of hard liquor (or a corresponding amount of alcohol) monthly or more often.

### Smoking

Smoking status was classified as daily smoking vs. other smoking statuses (including never, occasionally and quit smoking).

#### Sociodemographic predictors

Sociodemographic variables included sex, family structure and parental socioeconomic position (SEP) at age 16. Family structure was categorized into i) living with mother and father, ii) living with a parent and a stepparent and iii) other. Parental SEP was based occupation of parents (in TAM participants’ self-reports and in FinnTwin16 parents’ self-reports). SEP was classified into low (manual work), intermediate (lower nonmanual work) and high (upper nonmanual work) occupational position. If the occupation of neither parent was available, parental education was used and categorized as low (no occupational education), intermediate or high.

### Statistical analysis

Mplus Version 8.7 was used to obtain the joint trajectories of HED, smoking status and psychological distress. The participants were grouped using latent class analysis (LCA) based on their longitudinal profiles of HED, smoking and distress at ages 16, 22, 32, 42 and 52 (TAM) and 16, 25, 35 (FinnTwin16). The models were run for 1-8 classes. The Akaike information criterion (AIC), Bayesian information criterion (BIC), sample size adjusted BIC (ssaBIC), and Lo-Mendell-Rubin likelihood ratio test (LMR) were used as the statistical criteria to determine the optimal number of classes. Emphasis was also placed on group size (>5%) and interpretation. Participants were assigned to the latent classes according to most likely group membership based on estimated posterior probabilities.

For the LCA, we included those participants who had responded to questions about HED, smoking and distress each in at least two of the study waves (TAM, N=1943), (FinnTwin16, N=5166). To deal with missing values due to attrition we used the full information maximum likelihood estimation method. The nature of the twin data, i.e., within-pair/family-level clustering, was accounted for in LCA using the Mplus cluster option.

Finally, the associations between sociodemographic predictors and trajectory group membership were analysed using multinomial regression analysis. In the multinomial regression analysis, a generalized linear mixture model framework was used to account for family-level clustering of the FinnTwin16 data.

Two sets of sensitivity analyses were performed. First, to account for different follow-up times between the cohorts, we repeated the analyses harmonizing the follow-up times to resemble the cohort with shorter follow-up (i.e., FinnTwin16) (in the TAM, we included only ages 16, 22 and 32). Second, trajectory analyses were also performed stratified by sex due to possible known differences in the level and prevalence of symptoms and substance use between women and men.

## Results

The descriptive statistics of the study variables are presented in Table 1 and Appendix Figures 1-3. The means of psychological distress decreased from adolescence to adulthood in participants in the FinnTwin16 study and increased in the TAM study, with a peak at age 32. The frequency of HED in adolescence varied between 12.4% and 23.0% for women and between 13.9% and 27.5% for men. In FinnTwin16, the HED frequencies peaked at age 25 and subsequently decreased, whereas in women in the TAM there was a decrease at age 22, after which HED increased at age 32 and subsequently stabilized. In men in the TAM, HED increased until age 42. Daily smoking was most common in early adulthood (22/25 years, 23.5-33.9%) and decreased at later ages in both cohorts. Regarding sociodemographic variables, participants in the FinnTwin16 cohort had less often low socioeconomic background (24%) than those in the TAM cohort (50%).

**Table 1.** Prevalence of study variables among women and men in the TAM and FinnTwin16 cohorts.

| Variables | TAM |  |  | FinnTwin16 |  |  |
| --- | --- | --- | --- | --- | --- | --- |
|  | Women<br>Mean (SD) | Men<br>Mean (SD) | Total<br>Mean (SD) | Women<br>Mean (SD) | Men<br>Mean (SD) | Total<br>Mean (SD) |
| Psychological distress<br>age 16<sup>a, b</sup> | 4.6 (2.7) | 3.5 (2.7) | 4.1 (2.8) | 4.0 (2.6) | 3.2 (2.5) | 3.6 (2.9) |
| Psychological distress<br>age 22/25<sup>a, c</sup> | 4.7 (2.8) | 3.5 (2.9) | 4.1 (2.9) | 2.3 (2.9) | 1.8 (2.3) | 1.9 (2.7) |
| Psychological distress<br>age 32/35<sup>a, c</sup> | 5.4 (2.9) | 4.3 (3.1) | 4.9 (3.1) | 2.2 (3.1) | 1.6 (2.7) | 2.0 (2.9) |
| Psychological distress<br>age 42<sup>a</sup> | 5.0 (3.0) | 3.8 (3.1) | 4.5 (3.1) |  |  |  |
| Psychological distress<br>age 52<sup>a</sup> | 5.0 (3.2) | 4.1 (3.2) | 4.6 (3.3) |  |  |  |
|  | % (n) | % (n) | % (n) | % (n) | % (n) | % (n) |
| HED<sup>d</sup> age 16 | 23.0 (245) | 27.5 (307) | 25.3 (552) | 12.4 (370) | 13.9 (384) | 13.1 (754) |
| HED age 22/25 | 13.1 (116) | 29.6 (226) | 20.8 (342) | 41.7 (1174) | 68.2 (1647) | 53.9 (2821) |
| HED age 32/35 | 18.2 (146) | 51.1 (339) | 33.1 (485) | 20.6 (499) | 52.0 (1008) | 34.5 (1507) |
| HED age 42 | 18.5 (134) | 45.6 (272) | 30.7 (406) |  |  |  |
| HED age 52 | 16.6 (107) | 41.6 (212) | 27.7 (319) |  |  |  |
| Smoking age 16 | 19.4 (207) | 25.1 (281) | 22.3 (488) | 16.3 (483) | 19.3 (525) | 17.7 (1008) |
| Smoking age 22/25 | 27.5 (243) | 33.9 (259) | 30.5 (502) | 23.5 (640) | 32.5 (760) | 27.7 (1400) |
| Smoking age 32/35 | 20.0 (161) | 25.8 (171) | 22.7 (332) | 15.1 (353) | 21.1 (392) | 17.8 (745) |
| Smoking age 42 | 17.0 (125) | 22.1 (132) | 19.3 (257) |  |  |  |
| Smoking age 52 | 13.0 (84) | 13.9 (70) | 13.4 (154) |  |  |  |
| Parental socioeconomic<br>position age 16 |  |  |  |  |  |  |
| Upper nonmanual | 17.9 (188) | 19.5 (213) | 18.7 (401) | 35.3 (966) | 33.9 (876) | 34.6 (1842) |
| Lower nonmanual | 31.6 (332) | 30.8 (336) | 31.2 (668) | 41.0 (1123) | 41.0 (1059) | 41.0 (2182) |
| Manual | 50.5 (531) | 49.7 (543) | 50.1 (1074) | 23.7 (650) | 25.0 (646) | 24.4 (1296) |
| Family structure age 16 |  |  |  |  |  |  |
| Mother + father | 72.5 (738) | 75.5 (794) | 74.0 (1532) | 80.4 (2397) | 81.5 (2253) | 89.9 (4650) |
| Parent + step parent | 8.5 (87) | 8.9 (94) | 8.7 (181) | 6.6 (196) | 5.7 (158) | 6.2 (354) |
| Other | 19.0 (193) | 15.6 (164) | 17.2 (357) | 13.1 (390) | 12.8 (355) | 13.0 (745) |

<sup>a</sup> TAM: 7-item scale (range 0-21)
<sup>b</sup> FinnTwin16: 4-item scale (range 0-12)
<sup>c</sup> FinnTwin16: GHQ-12 (range 0-12)
<sup>d</sup> Heavy episodic drinking

Joint trajectories of HED, smoking status and psychological distress were identified using latent class analyses. In both cohorts, the statistical criteria e.g., the BIC, improved (decreased) continuously as the number of groups increased; however, increasing the number of classes decreased the size of the smallest class (Appendix Figure 4 and Appendix Table 1). Overall, the five class solution for the TAM and the three-class solution for FinnTwin16 were selected because they showed good fit indices and reasonable group sizes (<5%)(Appendix Table 1). The solutions are illustrated in Figure 1 and Figure 2.

**Figure 1.**
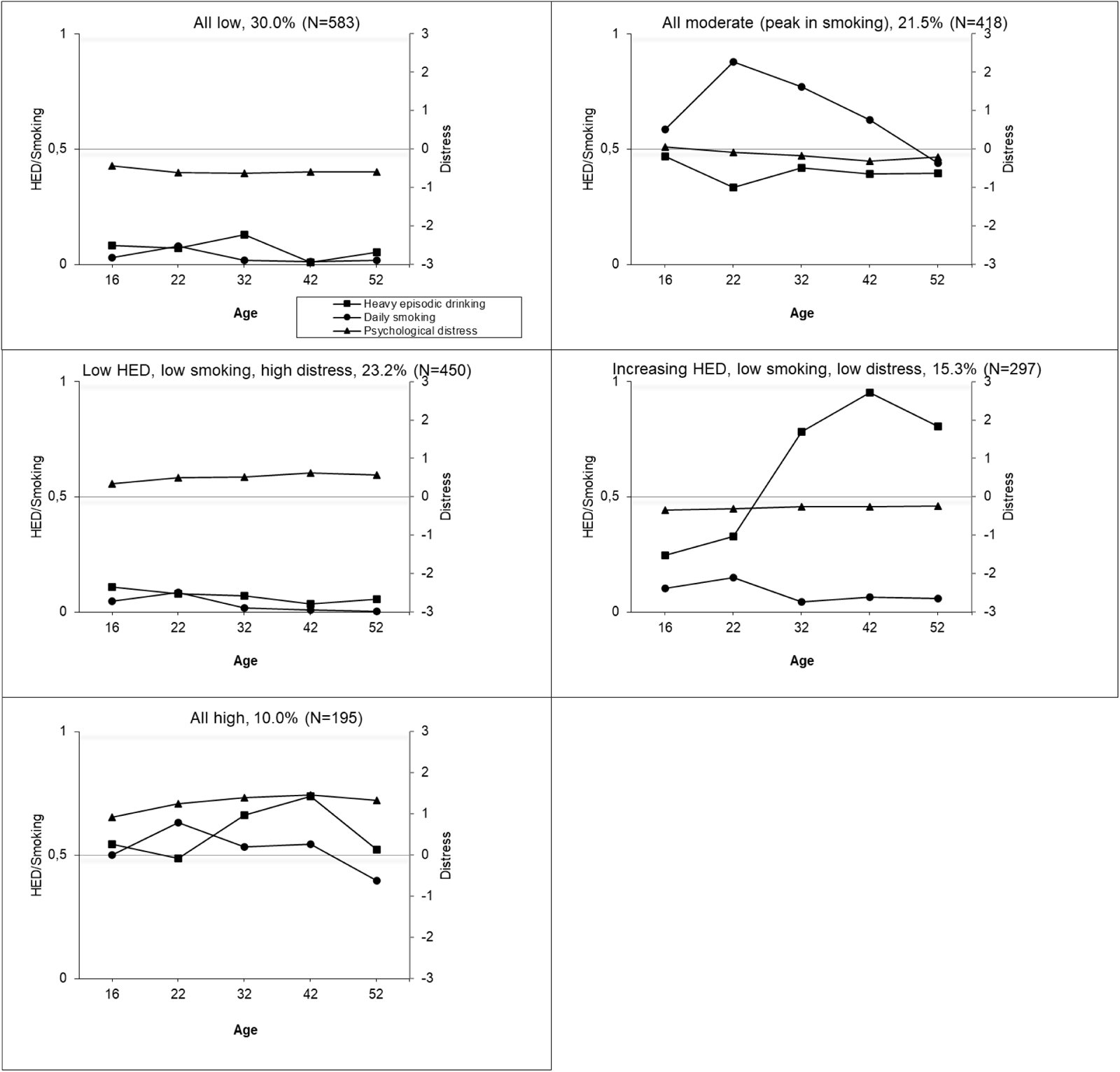
Conjoint trajectories of heavy episodic drinking, daily smoking and psychological distress among the TAM cohort at ages 16 to 52 years by a) class 1, b) class 2, c) class 3, etc. Score for HED: 0=less than monthly, 1= monthly or more often; for smoking: 0= less than daily, 1=daily smoking; for psychological distress: standardized.

**Figure 2.**
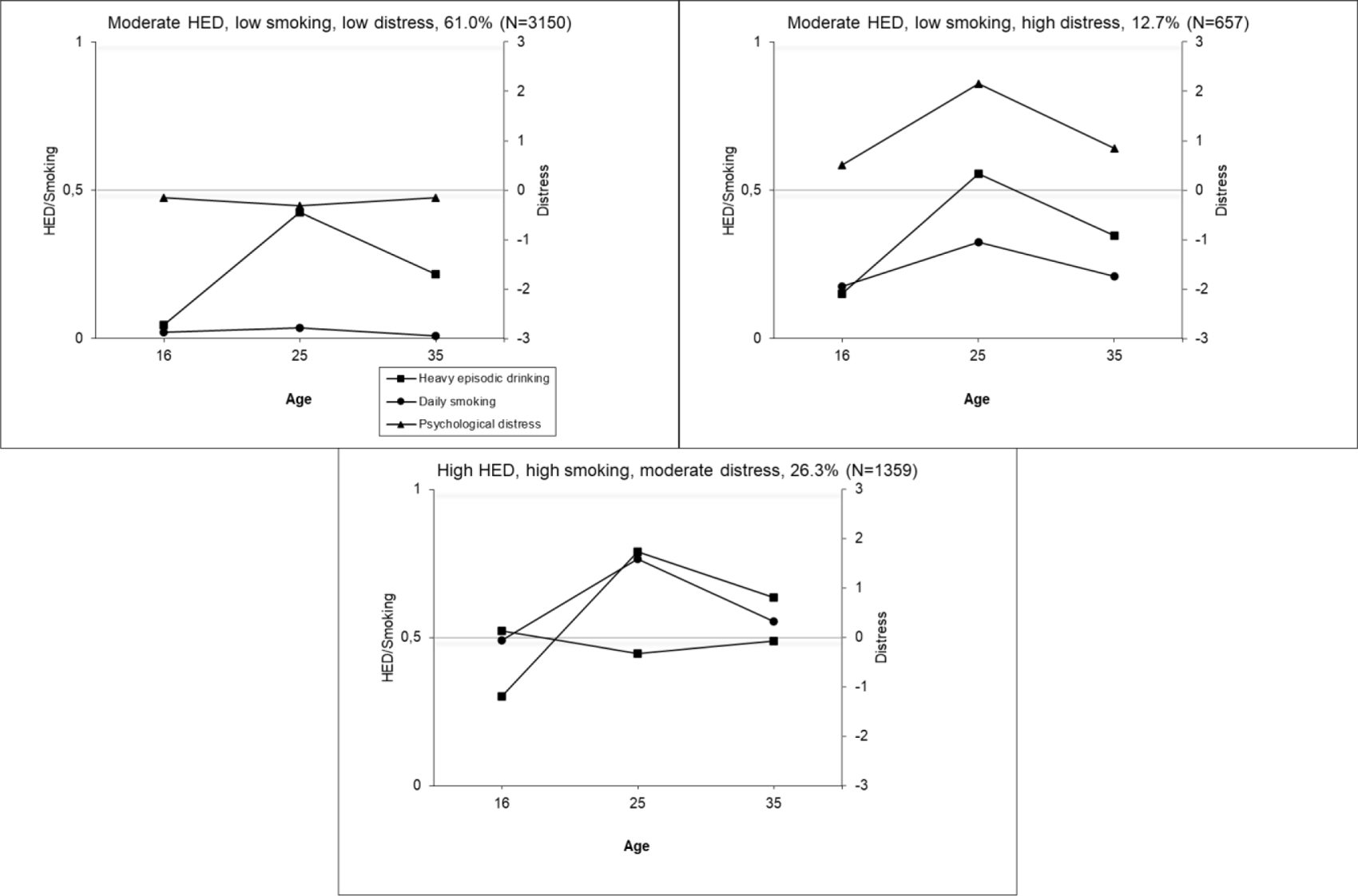
Conjoint trajectories of heavy episodic drinking, daily smoking and psychological distress among the FinnTwin16 cohort at ages 16 to 35 years by a) class 1, b) class 2, c) class 3. Score for HED: 0=less than monthly, 1= monthly or more often; for smoking: 0= less than daily, 1=daily smoking; for psychological distress: standardized.

The associations between sociodemographic variables and latent classes were analysed using multinomial regression analysis (Table 2). In the TAM men were more often assigned to trajectory groups all moderate and increasing HED, low smoking, low distress. Women were more often assigned to the trajectory group with low HED, low smoking, high distress. In FinnTwin16, men were more likely assigned to the high HED, high smoking, moderate distress group and women to the moderate HED, low smoking, high distress group. In general, in both cohorts, participants from families other than nuclear families were more often assigned to trajectory groups indicating high or moderate levels of substance use or distress compared to the group with low levels of use or distress. In the TAM, low parental SEP was associated with a likelihood of belonging to trajectory groups of i) all moderate, ii) low HED, low smoking, high distress, and iii) all high.

In Fintwin16, low parental SEP was associated with being more likely assigned to the high HED, high smoking, moderate distress group.

**Table 2.** Sociodemographic predictors of joint trajectories of heavy episodic drinking, smoking and psychological distress^a^.

|  | RRR (95% CI) | RRR (95% CI) | RRR (95% CI) | RRR (95% CI) |
| --- | --- | --- | --- | --- |
| **TAM<sup>b</sup>** | All moderate | Low HED, low smoking, high distress | Increasing HED, low smoking, low distress | All high |
| Sex (male) | 1.35 (1.05-1.74) | 0.33 (0.25-0.43) | 3.06 (2.25-4.16) | 1.00 (0.72-1.38) |
| Family structure |  |  |  |  |
| Mother + father | ref | ref | ref | ref |
| Parent + stepparent | 4.51 (2.69-7.56) | 2.04 (1.16-3.57) | 2.32 (1.27-4.22) | 6.73 (3.75-12.06) |
| Other | 1.8 (1.27-2.56) | 1.31 (0.92-1.86) | 1.08 (0.71-1.63) | 2.76 (1.81-4.22) |
| Parental SEP |  |  |  |  |
| Upper nonmanual | ref | ref | ref | ref |
| Lower nonmanual | 1.33 (0.91-1.96) | 1.20 (0.80-1.75) | 1.19 (0.80-1.75) | 1.10 (0.67-1.81) |
| Manual | 2.38 (1.67-3.40) | 1.51 (1.09-2.10) | 1.35 (0.93-1.95) | 2.01 (1.28-3.15) |
| **FinnTwin16<sup>c</sup>** | Moderate HED, low smoking, high distress | High HED, high smoking, moderate distress |  |  |
| Sex (male) | .54 (.45-.65) | 1.74 (1.51-2.01) |  |  |
| Family structure |  |  |  |  |
| Mother+father | ref | ref |  |  |
| Parent+stepparent | 1.41 (0.96-2.08) | 2.53 (1.88-3.40) |  |  |
| Other | 1.75 (1.36-2.25) | 1.84 (1.48-2.30) |  |  |
| Parental SEP |  |  |  |  |
| Upper nonmanual | ref | ref |  |  |
| Lower nonmanual | 0.87 (0.71-1.07) | 1.34 (1.11-1.61) |  |  |
| Manual | 0.88 (0.71-1.07) | 1.53 (1.24-1.89) |  |  |

<sup>a</sup> Relative risk ratios (RRRs) from multinomial logistic regression models (generalized linear mixture model for FinnTwin16). Bivariate models with only one predictor and trajectory group in the model at a time.
<sup>b</sup> Reference group All low.
<sup>c</sup> Reference group Moderate HED, low smoking, low distress.

## Discussion

The present study is one of the first to examine the joint trajectories of alcohol and tobacco use and mental health over the life course and the nature of their co-occurrence. The findings highlight that despite high correlations between these three outcomes, there is significant variation in how they jointly developed over time. In both examined cohorts, distinct groups of i) low levels of all three examined health concerns, ii) high levels of all three concerns, and iii) high distress and low-to-moderate substance use were found. In the older cohort with longer follow-up time, a group indicating iv) increasing HED and a group indicating v) all on average level, with a peak in smoking were also found.

Schulenberg et al (13) outline six interrelated components of a developmental perspective on substance use in the framework of LCT. These components can also be used in discussing the results of this study.

First, the *age curve* highlights the stability and change over time. Understanding the normative age curves of alcohol and tobacco use, as well as psychological distress, is essential for understanding their joint development. Despite heterogeneity in the number and timing of follow-ups as well as the measures, we found a well-acknowledged peak in substance use in early adulthood. In the older cohort (TAM), this was more evident regarding smoking. It has been shown in some previous Finnish studies that HED does not decrease after early adulthood especially among men, but rather increases until the mid-forties, which was also evident in the TAM cohort (56). Age curves of distress were rather stable in both cohorts and sexes, and it has been suggested that this difference in prevalence between women and men does not converge but continues into old age (43).

Second, there is *heterogeneity in the age curves,* as previous research has repeatedly shown, including previous studies with these two cohorts (38,57). What these previous studies have often lacked in their perspective is Schulenberg et al.’s third component of developmental perspective: *all else that is developing*. The development of HED does not occur in isolation, nor does the development of smoking behaviour or psychological distress. Some previous studies have attempted to address this challenge by examining the development of two phenomena at a time (32,33,38). This present study is one of the few studies that has tried to address this challenge even more broadly by exploring three developmental issues simultaneously. This study clearly shows that there are different joint patterns of HED, smoking and distress. The majority of people have low levels of substance use and distress symptoms during their life, and a minority continuously report high levels of all three of these health concerns. These groups indicate rather consistent developmental patterns from adolescence to adulthood. In this study, more than 60% of the younger cohort and approximately 30% of the older cohort were assigned to the group with consistently low levels of all three health concerns. In a similar study by Lee et al. (30) who followed participants from age 14 to 29, the proportion of participants in this group was comparable to that in the older cohort in this study (33%). In addition, the group indicating high/average levels through the follow-up was similar in Lee’s study and in our older cohort. In particular, adolescence and early adulthood are characterized by physiological and psychological changes, and the difference between normative development and poor health is not always clear (58). The cohorts studied here are population-based, and the overall age curves can be interpreted as normative development. By using trajectory analysis to examine these average curves in more detail, it is possible to distinguish normative and nonnormative development in these issues. The most problematic health concerns are those that continue to progress to later life stages (59,60). Overall, the trajectory groups showed minimal decreases in any of the three examined outcomes, especially in the older cohort with longer follow-up. However, in some groups, there were indications of the decrease in substance use after a peak in early adulthood.

Two interesting groups to consider are those with high distress symptoms: one with low and one with high levels of substance use. The identification and distinguishing of these groups (and their stable patterns) adds important information to mental health and substance use services. Long-lasting depressive and anxiety symptoms are important to treat, but substance use sets differing demands on clinical services (e.g. 61). From the public health perspective, it would be beneficial to focus on both alcohol use and smoking in mental health and substance use services, but often the substance in focus is alcohol (62–64). It has been shown that smoking dependence and smoking dependence motives related to heavy, automatic use and use to regulate affective states are associated with depression (65), highlighting the important independent role of smoking in these associations.

To our knowledge, trajectories of distress symptoms and alcohol and tobacco use have only been modelled jointly with three different datasets: the previously mentioned Harlem Longitudinal Development Study (30) and the cohorts used in this study. Although many similarities in the trajectory groups were found between these three cohorts, more studies with other cohorts should be conducted to be able to better differentiate the source of differences in the findings (population structure, follow-up time, drop-out, etc.). Future studies extending follow-up to older age groups (+50) are also important, as it has been shown that substance use is increasing in older age groups (66), and there is very little information on this rather new phenomenon, let alone its cooccurrence with mental health.

The fourth component of a developmental perspective considers a *larger sociocultural context* highlighting contexts structured by different ecological levels in individuals’ lives, ranging from socioeconomic characteristics to macrolevel societal policies and structures as well as historical time.

Both alcohol and tobacco are substances affecting health, but the development of their consumption in society has differed significantly in recent decades. HED has generally increased in Finland until the 2000s and then gradually started to decrease, although there are major socioeconomic disparities (67), while tobacco use has generally decreased (apart from snuff use) (CAN, 2017). The examined cohorts were born 7-12 years apart. The older cohort was in their twenties at the end of the 1980s, during the economic boom, and in their thirties during severe recession in Finland. The younger cohort was in their twenties and in the middle of the transition from school to work in the mid-1990s during the recession. A Swedish study revealed longitudinal differences between similar age cohorts in alcohol use but more so in total alcohol consumption than HED (68). A Finnish study that examined cohorts born between 1946 and 1977 found no cohort differences in alcohol use among men, whereas differences for women were found between every cohort; thus heavy drinking was more common for each new cohort than for the earlier cohorts, with the exception of the two youngest cohorts born after 1970, which resembled the younger cohort in this study (69).

The TAM cohort was obtained from one Finnish city, whereas FinnTwin16 is a national cohort. The TAM cohort was representative compared to the whole age cohort in Finland in terms of, for example, marital status (70) but was more educated. Compared to the population living in urban areas, the educational level was similar in the TAM cohort (71).

The fifth and sixth components embed the trajectory profiles into the full life course, the fifth highlighting the continuities and the sixth discontinuity. The fifth component concerns *long term developmental connections*, drawing attention to predictors and consequences of these trajectory groups. This study examined sex, family structure and parental SEP as potential predictors of trajectory groups. In general, male sex, other than nuclear family and low socioeconomic family background were associated with the probability of being assigned to a trajectory group other than the ‘all low’ trajectory group. As we hypothesized, men were more often assigned to groups with average/high substance use, and women were more often assigned to groups indicating high distress but low substance use. This sex difference has been evident in previous studies and has now also been verified in this study by modelling all three aspects simultaneously. In general, in the sensitivity analysis, the same trajectory groups were identified for men and women, but these sex stratified analyses also showed similar tendencies in sex differences. For example, in the TAM cohort in women in the high distress, low substance use group, the high level of distress was more pronounced than that in the whole group. In men, the group of increasing HED was more pronounced than the distress group. This sex difference was also shown in our analysis when pooling women and men, which justifies our decision to analyse combined data.

Future studies should aim to combine the third and fifth components of the developmental perspective: simultaneously model the development of several different health concerns and identify the precursors (and consequences) of these heterogenous longitudinal profiles. The question about predictors and consequences is also relevant regarding the trajectory groups per se. The aim of this paper was not to determine the causal paths behind these associations, but it should be acknowledged that the directions are likely reciprocal, and several underlying mechanisms may explain these associations. However, several studies have suggested that these associations between mental health and alcohol use exist at some level even after controlling for these kinds of confounders (3,72,73). Regarding smoking and mental health some studies have suggested that the association is confounded by shared genetics and other familial liabilities (74).

The sixth and final component of Schulenberg et al.’s developmental perspective highlights the *interplay between developmentally distal and proximal effects*. We know from previous single trajectory studies that not all substance use or mental health concerns start in adolescence or early adulthood. Moreover, not all early health problems reflect enduring difficulties. (e.g. 57). Early adulthood has typically been a life phase with a peak in distress and substance use. Many mental health problems initiate in adolescence/early adulthood and experimenting alcohol or smoking at that life stage may be a starting point for an addictive path. Identifying those who are at risk of developing long-term problems is essential. This places important demands on substance use and mental health services.

### Methodological considerations

This study used data from two large cohorts that followed participants several years prospectively through different life stages and had reasonably good participation rates. However, there was attrition in both cohorts. In the TAM cohort, when accounting for several age 16 variables simultaneously, male sex and poor school performance at age 16, but not substance use or distress, predicted a lower number of responses between ages 22 and 52 (range 0–4). (51). Nevertheless, there were, on average, 3.6 out of five responses per participant. In FinnTwin16 male sex and low parental SEP at age 16 predicted nonparticipation at age 35.

Future research should aim to study substance use and distress in more detail. In this study, depressive and anxiety symptoms were combined into a more general concept of psychological distress. In future studies, the development of these two scopes would also be useful to differentiate, as they may have somewhat differing associations with substance use. In addition, we did not measure the amount of tobacco used.

To address differences between the cohorts in follow-up times (36 vs. 19 years), sensitivity analyses were performed analysing TAM data at only the first three time points. These analyses identified trajectory groups similar to those identified when analysing all five time points. The only trajectory group missing using a shorter follow-up time was increased HED, low smoking status, and low distress. This finding indicates previously mentioned interesting findings typical of Finnish men’s drinking habits; HED does not decrease after early adulthood, but rather later in mid-adulthood (56,57). This highlights the importance of extending follow-up times beyond early adulthood when examining trajectories of substance use.

Although there are clear advantages of being able to explore study questions using multiple existing datasets, retrospective data harmonization is challenging. Within-study differences in variables over time and between-study differences should be acknowledged when interpreting the results of this study.

## Conclusions

This study identified several different patterns of how heavy episodic drinking, daily smoking and psychological distress cluster across time from adolescence to adulthood. The trajectory groups identified in this study add a new perspective to the well-known complexity of these associations. Despite the high correlation between these issues, they are often treated separately in the service sector (49). Our results highlight the importance of taking a more holistic approach, recognizing the heterogeneity in people with various combinations of history of health concerns and tailoring preventive and treatment interventions accordingly. Life phase-specific interventions are needed to prevent the development of risky substance use and poor mental health as both separate and cooccurring public health problems.

## List of abbreviations

AIC: Akaike information criterion
BIC: Bayesian Information Criteria
HED: heavy episodic drinking
LCA: latent class analysis
LCT: life course theory
LMR: Lo-Mendell-Rubin likelihood ratio test
RRR: relative risk ratio
ssaBIC: sample size adjusted BIC
SEP: socioeconomic position
TAM: the ‘Stress, development and mental health’ cohort

## Declarations

### Ethics approval and consent to participate

The TAM study was approved by the Ethics Committee of the Finnish Institute for Health and Welfare. FinnTwin16 was approved by the Ethical Committee of the Department of Public Health, University of Helsinki, Helsinki University Central Hospital ethical committee, and by the Institutional Review Board of Indiana University. All participants gave informed consent to participate.

### Consent for publication

Not applicable.

### Availability of data and materials

The information on the datasets analysed during the current study is available on the following webpages www.thl.fi/en/tam (TAM cohort) and https://wiki.helsinki.fi/xwiki/bin/view/twinstudy/Kaksostutkimus/

(Finntwin16). The data underlying this article cannot be shared publicly due to legal restrictions (Finnish Data Protection Act 1050/2018) and the nature of the data (individual level data).

The data are available upon request. Data requests are reviewed at the Finnish Institute for Health and Welfare (THL)(for TAM) and the Institute for Molecular Medicine Finland (FIMM) Data Access Committee (DAC) (for FinnTwin16) for authorized researchers who have IRB/ethics approval and an institutionally approved study plan. For more details, please contact THL and the FIMM DAC (fimm-dac@helsinki.f).

### Competing interests

The authors declare that they have no competing interests.

### Funding

This work was supported by the Research Council of Finland (Academy of Finland) (grant #339114), the Academy of Finland Center of Excellence in Complex Disease Genetics (grant #352792), the Finnish Foundation for Alcohol Studies and the Juho Vainio Foundation. None of the funders had any role in the design of the study, the collection, analysis, or interpretation of the data; or in the writing of the manuscript.

### Authors’ contributions

NB performed all the statistical analyses and NB, MP, AL, MM and OK interpreted the analyses. NB drafted the manuscript, which was revised by the remaining authors (MP, AL, MM and OK). All the authors have read and approved the final manuscript.

## Acknowledgements

Not applicable.

---

**Appendix Figure 1.**
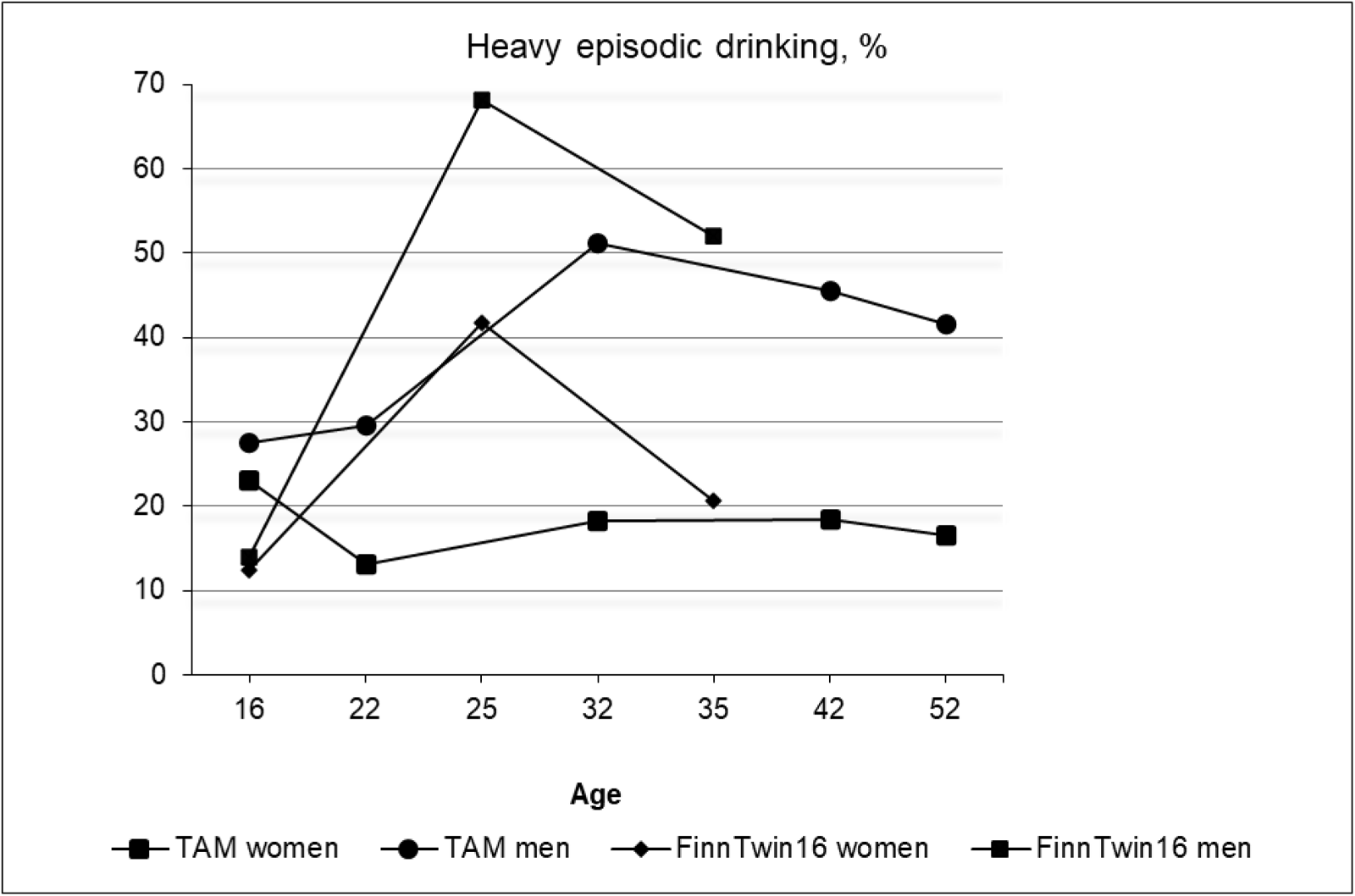
Frequencies of heavy episodic drinking in men and women in the TAM and FinnTwin16 cohorts

**Appendix Figure 2.**
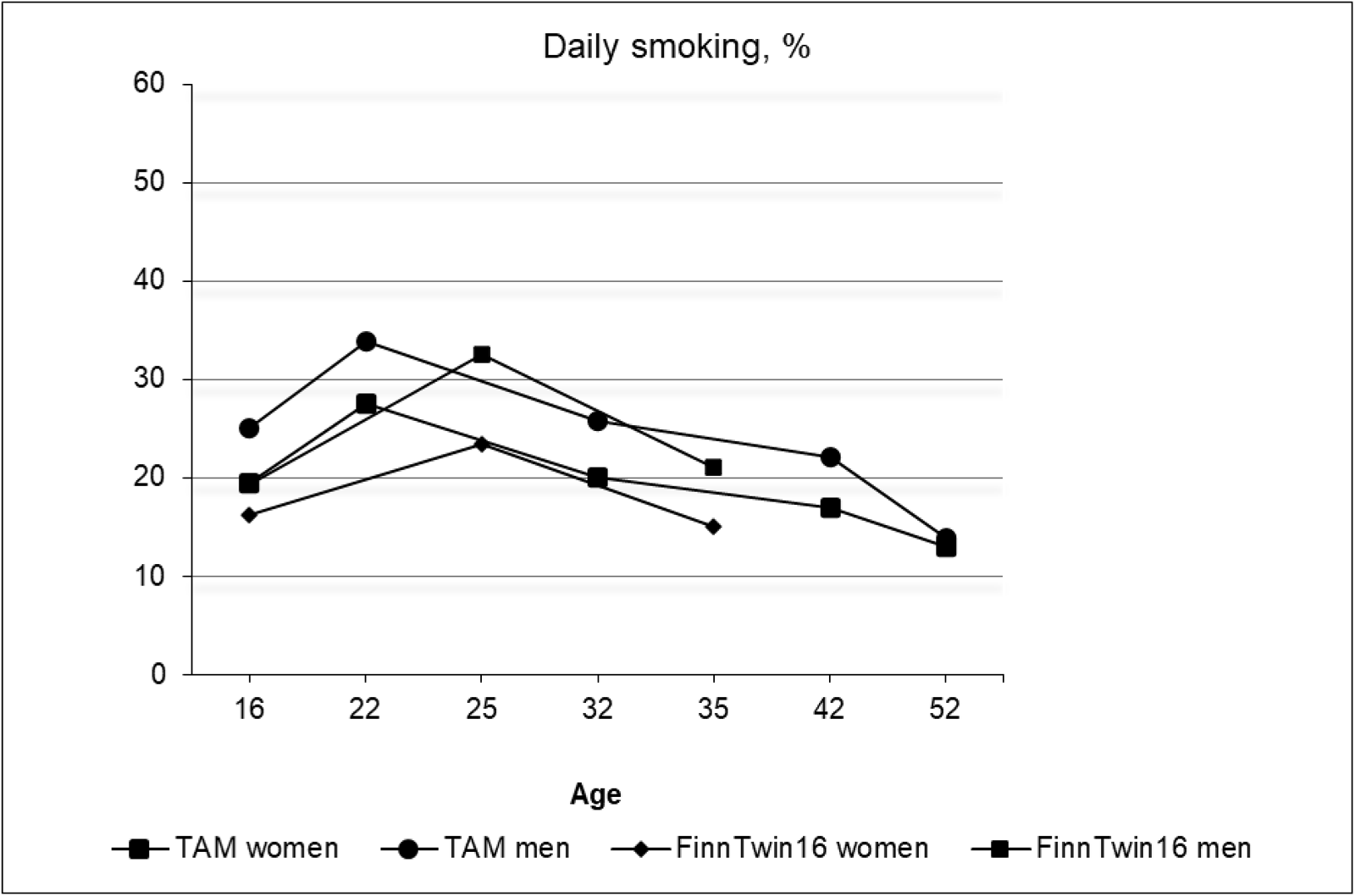
Frequencies of daily smoking in men and women in the TAM and FinnTwin16 cohorts

**Appendix Figure 3.**
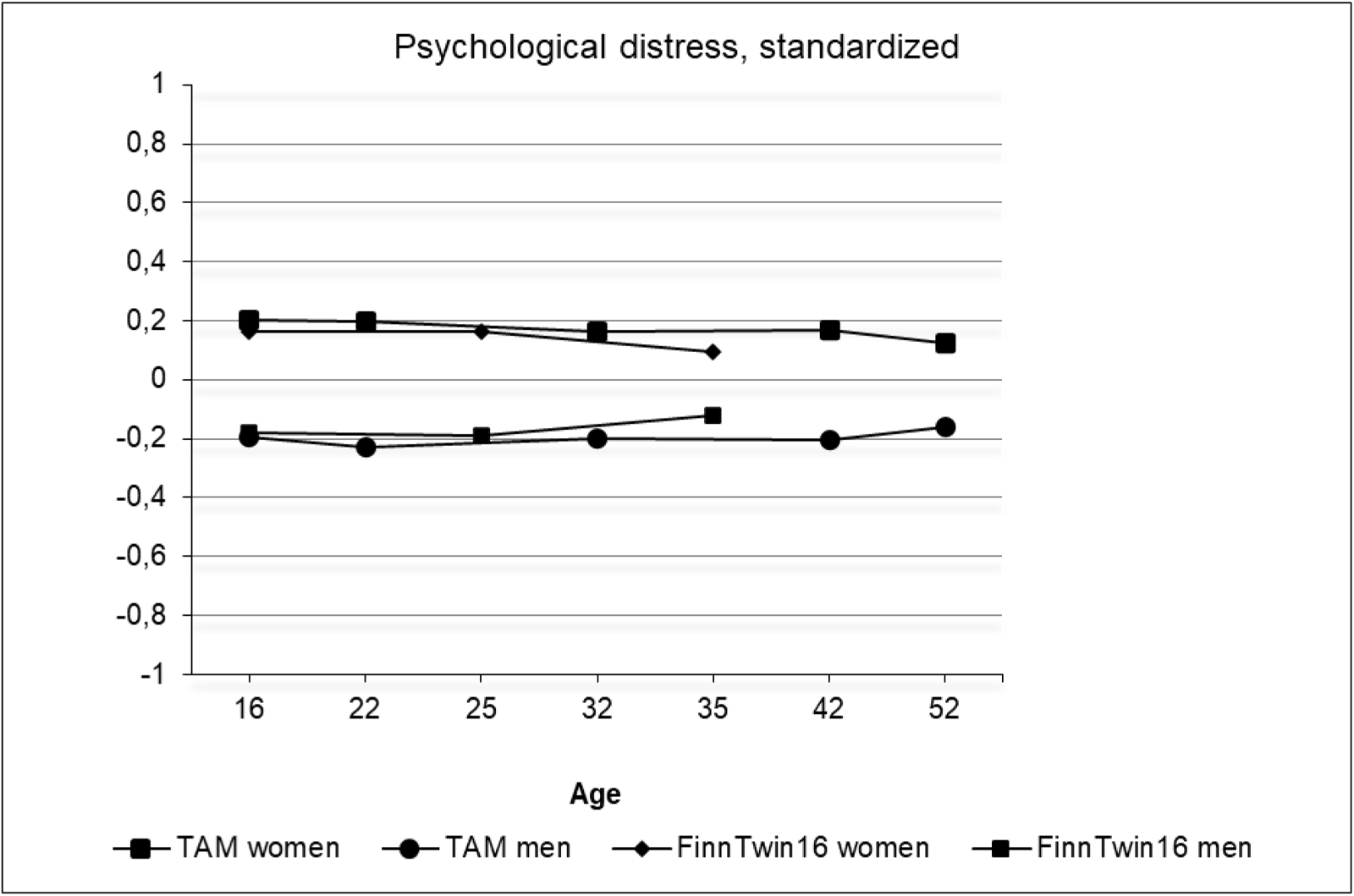
Standardized means of psychological distress in men and women in the TAM and FinnTwin16 cohorts

**Appendix Figure 4.**
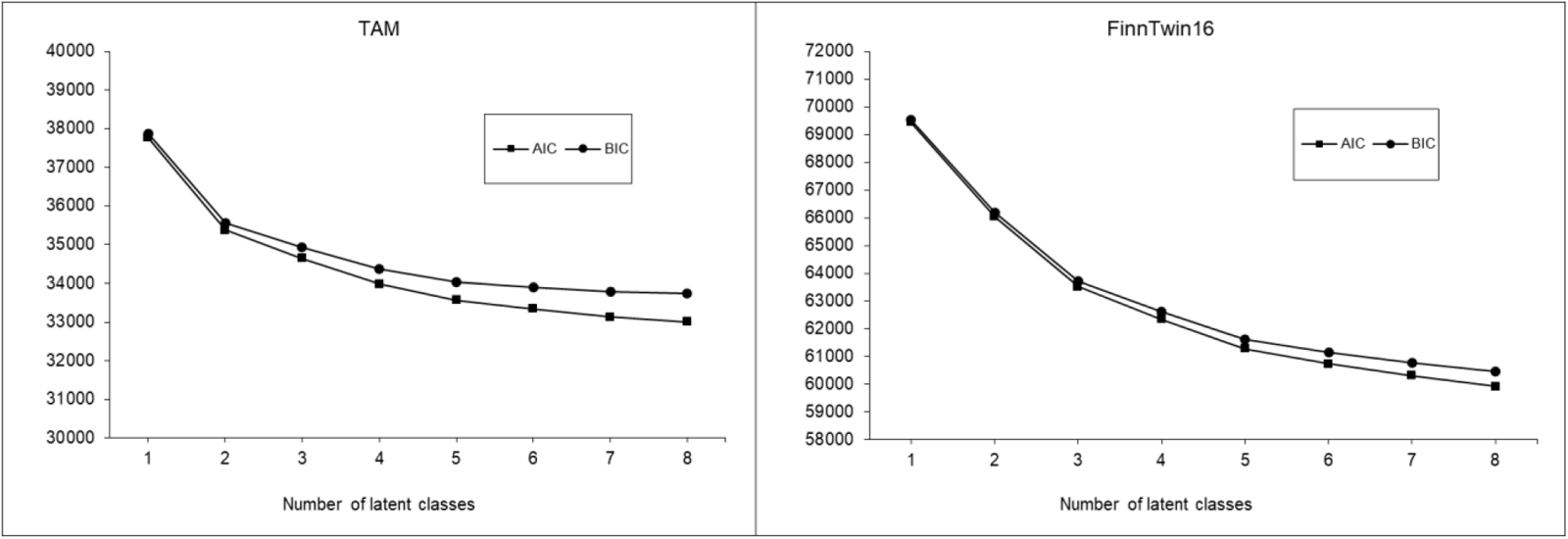
Fit statistics (AIC and BIC) by number of latent classes and cohorts

**Appendix Table 1.**
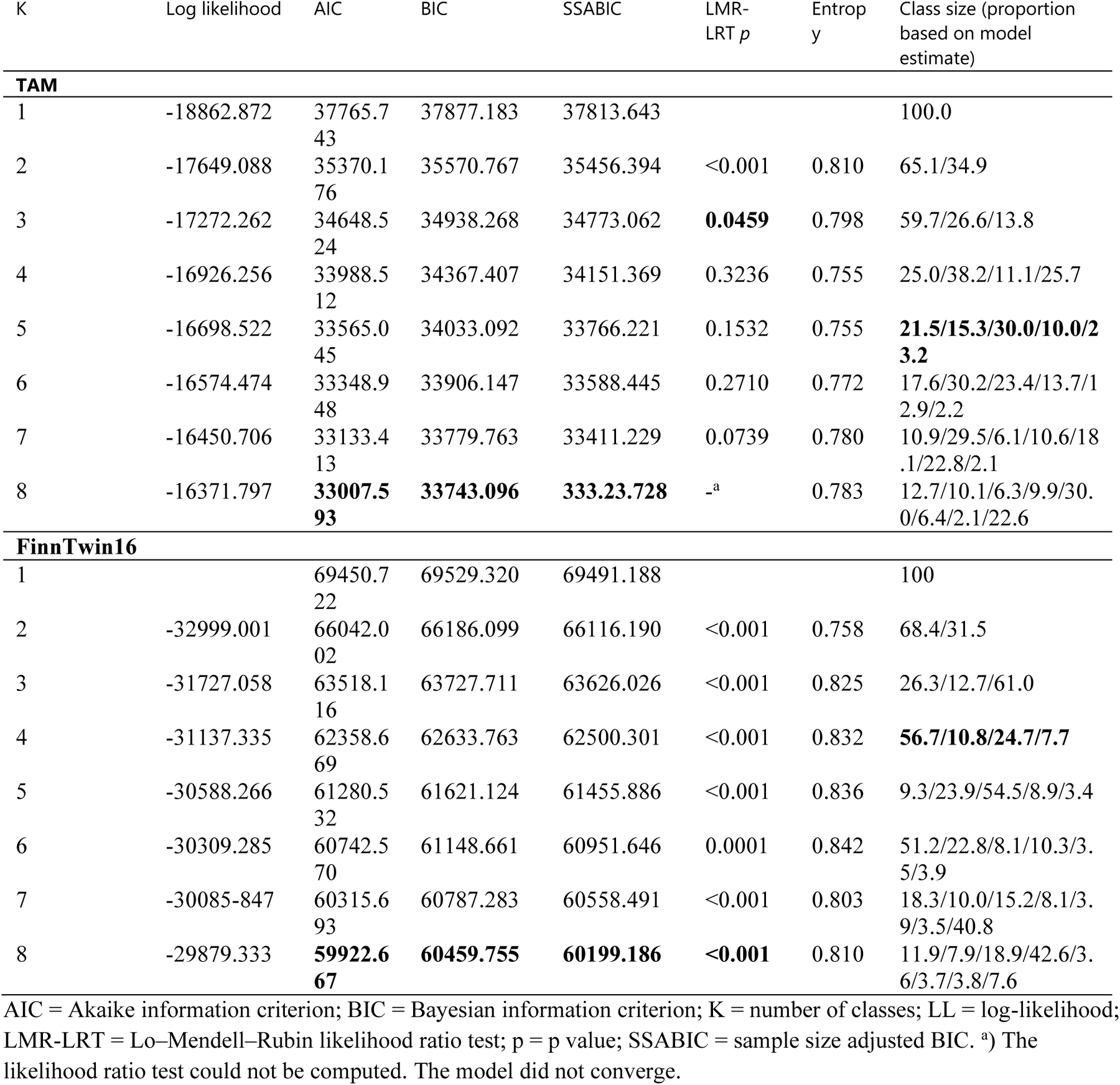
Fit statistics, classification indices, and class sizes for each joint trajectory model of heavy episodic drinking, smoking and psychological distress.

## Notes

### Competing Interest Statement

The authors have declared no competing interest.

### Author Declarations

Ethics committees/IRBs of 1) Finnish Instute for Health and Welfare, 2) Department of Public Health, University of Helsinki, 3) Helsinki University Central Hospital, and 4) Indiana University gave ethical approval for this work.

